# Theory-guided development of fertility care implementation strategies for adolescents and young adult cancer survivors

**DOI:** 10.1101/2020.06.30.20143891

**Authors:** Anna Dornish, Emily M. Yang, Jamie Gruspe, Erin R. Roesch, Paula Aristizabal, Greg A. Aarons, Sally A.D. Romero, Michelle B. Takemoto, Bonnie N. Kaiser, H. Irene Su

## Abstract

**Background:** Oncofertility care remains under-implemented across oncology and fertility care settings, with limited tools to scale up effective implementation strategies. Guided by the Consolidated Framework for Implementation Research (CFIR), we aimed to systematically assess factors that influence implementation of oncofertility care and map strategies, particularly electronic health record (EHR)-enabled ones, that fit adult and pediatric oncology care contexts.

**Methods:** Using purposeful sampling, we recruited healthcare providers and female adolescent and young adult (AYA) cancer survivors from a comprehensive cancer center and a freestanding children’s hospital. Participants underwent semi-structured interviews and focus groups. Using thematic analysis combining inductive codes with CFIR-based deductive codes, we characterized barriers and facilitators to oncofertility care and implementation strategies. Two coders independently coded each transcript, with a third coder resolving discrepancies by consensus.

**Results:** We recruited 19 oncology and fertility providers and 9 AYA survivors. We identified barriers and facilitators to fertility care in the CFIR domains of individual, inner setting, outer setting, and process, allowing us to conceptualize oncofertility care in three necessary stages: screening, referral, and fertility preservation counseling. To fit an adult and a children’s context, five implementation strategies were mapped: needs screen using a best practice advisory, referral order, telehealth fertility counseling, provider audit and feedback, and a provider educational session. All but provider education are facilitated by the EHR system.

**Conclusions:** An implementation science approach enabled systematic assessment of oncofertility care and co-design of implementation strategies with stakeholders, providing a theory-based approach and scalable EHR tools to support wider dissemination.

## Background

Unmet reproductive health care needs are highly prevalent among adolescent and young adult female cancer survivors (AYA survivors). In the U.S., there are nearly 360,000 AYA survivors who are younger than age 40.^1^ AYA survivors undergo treatments such as radiation, chemotherapy, surgery, and/or endocrine therapy in a variety of clinical settings, from comprehensive cancer centers to children’s hospitals. Such treatments may adversely impact future fertility.^2,3^ AYA survivors often want to be able to have their own families; infertility from cancer treatment is devastating and significantly impairs quality of life.^4,5^ However, fertility care – fertility counseling and fertility preservation procedures – at cancer diagnosis and post-treatment can prevent infertility.^6,7^ Hence, clinical guidelines from oncology and fertility societies recommend that oncologists should discuss the possibility of infertility with patients and offer patients fertility preservation options or referrals to reproductive specialists^8^ throughout the cancer continuum.

Despite clinical guidelines, uptake of fertility counseling remains highly variable. A 2015 report of four NCI-designated comprehensive cancer centers found that among reproductive age patients, only 26% received documented fertility counseling on infertility risks at cancer diagnosis, and 24% received fertility preservation options, with 13% documented referral to a fertility specialist.^9^ Low implementation of fertility counseling is attributed to heterogeneous barriers and facilitators.^10^ Examples include inadequate recognition of reproductive health needs by patients and providers,^11,12^ unclear role expectations of oncology versus fertility providers,^13- 16^ lack of clear referral pathways,^17-19^ and lack of access to fertility programs, particularly in pediatric oncology settings.^20^ A limitation of prior research is the lack of systematic approaches to assess barriers and facilitators in a health system and map scalable strategies to implement routine fertility counseling.

Electronic health records (EHRs) may facilitate implementation strategies that enable fertility counseling. EHR systems can set rule-based reminders to staff and/or providers, automate referral pathways, generate reports of fertility referral and counseling, collect patient-reported information through a patient portal and support telehealth. For widely used EHRs, functionalities are shareable, and mobile apps are easy to upgrade universally. Moreover, connectivity via smartphones is more than 80% of AYAs, regardless of socioeconomic status, facilitating reach.^21^

To date, an implementation science approach – the study of methods to promote integration of evidence-based practices into routine health care – has not been undertaken to address the know-do gap in fertility counseling.^22^ Thus, guided by the Consolidated Framework for Implementation Research (CFIR),^23^ we systematically assessed barriers and facilitators to fertility counseling and use of EHR tools as implementation strategies to integrate fertility counseling into two oncology programs, one adult and one pediatric. We compared adult versus pediatric and inpatient versus outpatient settings. We then designed a multi-component implementation strategy to fit the two clinical contexts.

## Methods

The study was approved by IRBs at University of California San Diego (UCSD) and Rady Children’s Hospital. Researchers were female oncologists, reproductive endocrinologists, implementation scientists, and medical students.

Between October 2018-May 2019, we enrolled reproductive-aged female AYA survivors and their healthcare providers to participate in semi-structured interviews or focus groups. Study settings were UCSD Moores Cancer Center, an adult comprehensive cancer clinic at a tertiary academic medical center, and Rady Children’s Hospital San Diego, a freestanding children’s hospital affiliated with UCSD. At Moores, disease teams included hematologic malignancies, gastrointestinal, neuro-oncology, breast oncology, and radiation oncology. At Rady Children’s, disease teams included liquid tumor, solid tumor, bone marrow transplant, and the survivorship clinic.

We developed CFIR-based guides for provider interviews and AYA survivor focus groups to assess barriers and facilitators to fertility counseling as well as explore use of EHR-based implementation strategies into routine oncology care. The guides encompassed questions on the 5 CFIR domains (intervention, individual, inner setting, outer setting, process) and relevant domain-specific constructs. CFIR was selected for its emphasis on multilevel ecological factors, which would enable both a systematic assessment as well as mapping implementation strategies by level and construct.^23^

We conducted 19 health care provider semi-structured interviews, 8 at the adult program and 11 at the pediatric program. Oncologists and advanced practice providers from each disease team were approached for participation, because clinic processes and complement varied by disease team. Program clinical and quality leaders were also recruited. Informed consent was reviewed with participants prior to the interview. Interviews were 1-hour long either in person or via video calls of the participant with 2 or 3 investigators.

We conducted 4 focus groups with 2-3 reproductive-aged AYA survivor participants per group. They were recruited from the investigators’ prior research studies on reproductive health in AYA survivors.^24,25^ Among participants who agreed to be contacted for future studies, we restricted to individuals who received care at either of the two oncology programs and were younger than 45. Participants received recruitment emails, questions were answered by the study team, and consents were signed and returned prior to video focus groups. Focus groups were 1-hour long via video calls with 2-3 investigators.

Audio recording and note-taking took place during interviews and focus groups. Participants were compensated with e-giftcards. Recruitment was stopped when data saturation was achieved.

### Data analysis

We conducted thematic analysis facilitated by MaxQDA software. ^26^ In addition to deductive themes (e.g., CFIR constructs ^23, 27^), we identified inductive themes, or those arising from the data, using the following steps: 1) two independent coders (AD, JG) read the transcripts, becoming familiar with the text and developing initial codes by consensus, 2) they coded three transcripts iteratively and refined the codebook, 3) the final codebook was determined by consensus (AD, JG, HIS), 4) all data were coded, and 5) data were summarized by theme, with systematic comparison of pediatric versus adult settings and inpatient versus outpatient settings.^28^

## Results

Ten physicians (8 medical oncologists, 1 surgical oncologist, and 1 radiation oncologist), 2 advanced practice providers, 1 pharmacist, and 6 nurses participated. Our sample had 8 adult providers and 11 pediatric providers. There were 16 women and 3 men. Among the nine AYA survivors, mean age was 33.1 (SD 6.8) years, and their cancer diagnoses included thyroid, cervical and bone cancers, leukemia and lymphomas. Figure 1 summarizes barriers and facilitators to fertility counseling by CFIR domains (i.e., individual characteristics, inner context, outer context, process) and related constructs.

**Figure 1:**
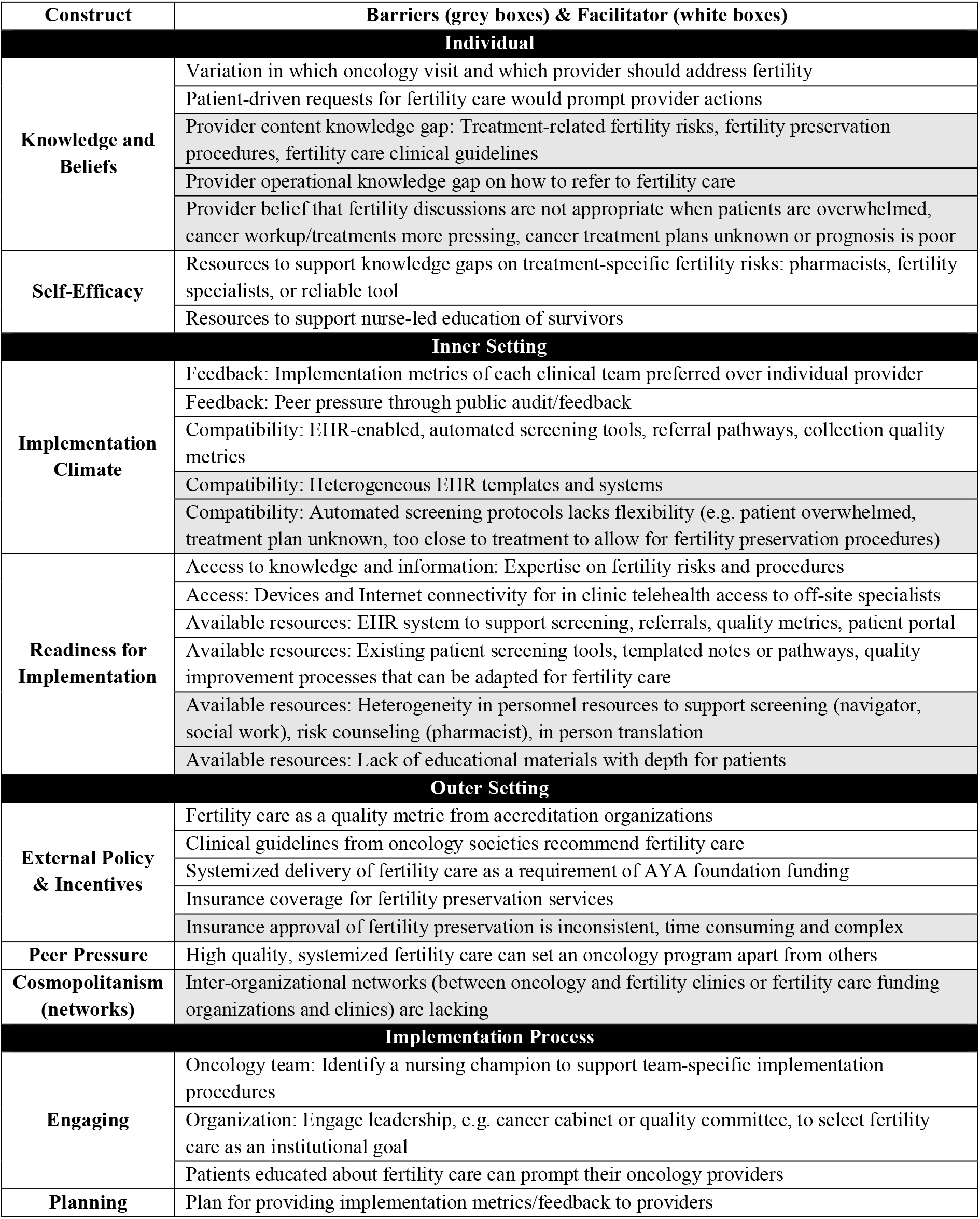
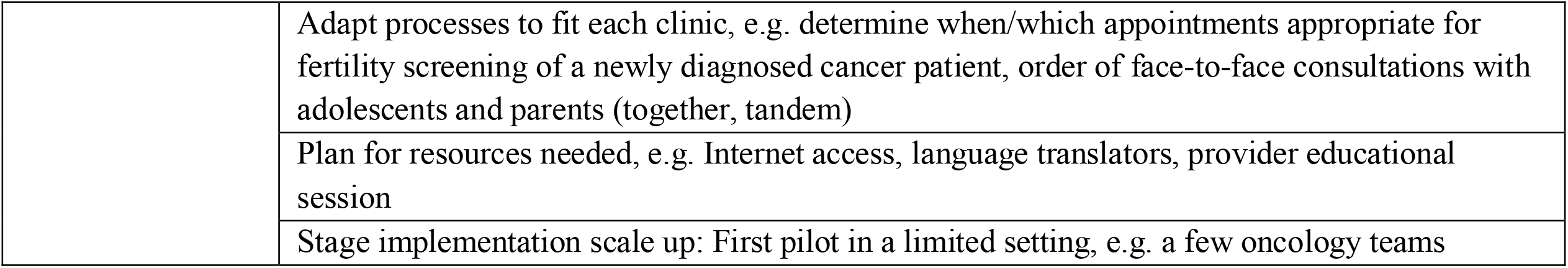
Barriers (grey boxes) and facilitators (white boxes) in implementation of fertility care by CFIR domains and constructs

### Individual characteristics

Providers had variable content knowledge about the existence of fertility care guidelines, fertility risks of cancer treatments, and fertility preservation procedures. Even for oncology providers who addressed fertility, depth of content knowledge was sometimes perceived to be lacking: “*[I] feel as though as I am inadequately trained in this arena. Having said all of that, I do feel like we are addressing the issue and at least putting it on the table, but I don’t think we’re doing enough*” (Advanced practice provider). This content knowledge gap limited self-efficacy; access to knowledge on fertility risks, which may be derived from a number of resources, was felt to be key to improving self-efficacy. Beyond content knowledge, oncology providers lacked knowledge on how fertility counseling and referral are operationalized in their clinic.

From the perspective of AYA survivors, fertility counseling would inform their decisions about timing of cancer treatment, and lack of knowledge about the option of fertility preservation procedures prevented AYA survivors from undertaking them:

> *I think before I would have liked to know that. In my opinion, I think doctors being doctors push, push treatment right away. And if I had known, like … if I would’ve been able to preserve or do something for fertility, I think I would’ve chosen to wait on treatment and done that. – AYA survivor*

There was also variation in beliefs on when fertility counseling is appropriate and who should provide the counseling. While nearly all providers expressed belief that fertility care is relevant to AYA survivors, the timing of counseling at cancer diagnosis can be complicated by overwhelmed patients, competing oncology workup and/or treatment initiation needs, unclear cancer treatment plans that preclude informing patients about their reproductive risks, and poor prognosis: “*I don’t know if they’re going to need chemo because for various reasons, and they’re going to surgery first, so I don’t bring it up at all. And for the patients that I’m sending for neoadjuvant, I just assume the oncologist is going to do that unless they ask me specifically. If the patient asks me about it then I’ll put the referral in”* (Surgical oncologist). As suggested by this oncologist, regardless of if/when a provider brings up fertility, patient-driven requests are a useful cue to action.

Preferences varied on which provider – i.e., physician (oncology or fertility), advanced practice providers, or nursing – would conduct the primary fertility counseling. The oncology physicians reporting self-efficacy prefer to undertake primary counseling themselves, while others would opt for automated referrals for all survivors to fertility specialists. There was consensus that nursing has an important role in patient education, inclusive of the potential for fertility education, but this requires significant education and materials nurses can use for teaching.

### Inner context

Discussions of characteristics of the clinic settings important to delivering fertility counseling focused on the implementation climate and readiness. EHR tools were suggested and/or endorsed by providers as compatible with automating screening for fertility needs in clinic or before visits via patient portal, referral pathways between oncology and fertility, and accrual of fertility care metrics. Advantages were alleviating personnel workload, systematically selecting the targeted population (e.g. via setting age parameters and type of encounter for automated fertility needs screens) and generating shareable tools among organizations using the same EHR platform. EHR documentation, particularly with discrete fields, allows efficient collection of quality metrics for feedback and accreditation: “*It’s also important to have the resource of being able to pull metrics because otherwise you have this warm fuzzy feeling in your heart that we’re doing super well, but then the data shows that that was an erroneous warm fuzzy feeling”* (Quality leader). Providers had experienced feedback of quality metrics as a clinic team and individually through quality improvement initiatives and preferred team-based feedback.

Participants emphasized that automated EHR pathways still require human action, and personnel resources varied by clinic teams. When considering a health screen entered into the EHR by medical assistants, a physician stated that they were “*not aware of what those answers are when [seeing the patient] at the first appointment. So that’s the problem with the digital ones, is that the [medical assistant] might collect the information, put it in at the end of the day, and then I’ll see it in EPIC after I’ve seen [the patient]… if they could verbally flag, that’s probably the best way to inform clinicians*.*”* Personnel resources to support EHR tools varied but included patient navigators, patient access representatives to help patients register for patient portals, and in-person, telephone- or video-based language translators. Neither oncology program had institutional resources supporting a dedicated fertility navigator.

The third theme centered on adapting available resources to facilitate use, including paper or EHR screening tools, EHR note templates, quality metrics reporting, and quality improvement processes for fertility counseling implementation. In addition, all levels of providers noted a lack of fertility educational materials that were sufficiently in-depth and had patient-friendly content: “*Recently, for example, I had a patient [… and] we didn’t have the resources here. We had to try and pull resources and then go and discuss with the patients. I don’t think as pediatric oncologists we have been trained to do that*.*”* An oncology nurse echoed, *“I mean fertility is such a sensitive topic for families, when they hear that, it just kind of like breaks their heart. So we wouldn’t want to… we would want to make sure that if we start talking about it, we have the appropriate resources for them*.*”*

### Outer context

External factors influence clinic and individual providers’ delivery of fertility care. An oncology physician noted, *“[At] national meetings [oncofertility] is always a big topic, something that’s on our radar a lot*.*”* Additionally, advocacy and funding organizations use fertility care as a quality metric as well as a requirement for receiving funding:

> *One of [a pediatric oncology non-profit’s] big requirements is to have this fertility preservation talk at the very beginning, before starting chemotherapy. So, we have to come up with some kind of protocol to put them into our program where it just happens all the time… They require that to get their sponsorship, we have to have formal protocol in place. –* Pediatric oncologist

Insurance coverage for fertility preservation services was repeatedly discussed as a barrier. Even if insurance coverage for fertility counseling is available to patients, procedures may not be covered and authorization processes are burdensome. As an example, after a fertility preservation referral order has been placed in one oncology program, insurance authorization is required and obtained by a central or provider team-specific authorization unit. The authorization comes as a physical letter, and to bypass the delay, the provider team makes additional calls to make sure their referrals are getting authorized.

Lastly, delivery of fertility care is complex due to involvement of different organizations, i.e., oncology clinics, fertility clinics, non-profit organizations, and insurance companies. Oncology programs do not always have a fertility program in the same healthcare institution. Participants noted a strong need to bridge these different organizations systematically and seamlessly.

Oncology and fertility clinics also need to be part of a network for fertility-related financial support from non-profit organizations such as Livestrong. Integration and the resulting interconnectedness between organizations needs to be systematic.

### Process

Many providers reflected on the types of engagement needed for effective implementation. Leadership engagement at three distinct levels were proposed: organizational level that sets fertility care as an institutional goal (e.g., selection of the Quality Oncology Practice Initiative (QOPI) quality metric ^29^), cancer team leaders that endorse care goals for the team, and a fertility champion within each team. Largely, nurses were identified as the ideal fertility champion within teams to increase engagement. Providers viewed engagement of multiple people as a facilitator because *“people feel resistance when it’s like this is one person’s project”* (Oncologist). Moreover, engaging AYA survivors as their own advocates would effectively cue their providers.

Co-planning implementation strategies with stakeholders was endorsed as important for fit and buy-in. Adaptation of processes will be needed to fit clinic context, e.g. which appointments are appropriate for introducing fertility needs screens and whether AYA survivors are seen in sequence or concomitantly with their parents. Oncology providers also desired input on which fertility care metrics should be provided to whom as feedback. Lastly, several providers suggested piloting implementation strategies with engaged teams first to identify problems before full-scale implementation.

Three stages emerged as important to planning fertility counseling implementation (Figure 2). First, patients need to be screened for fertility care needs. Second, a fertility care referral needs to be placed, if appropriate. Third, patients need access to a fertility specialist to undergo additional fertility counseling and appropriate fertility preservation strategies. Different types of individuals can participate in each stage.

**Figure 2:**
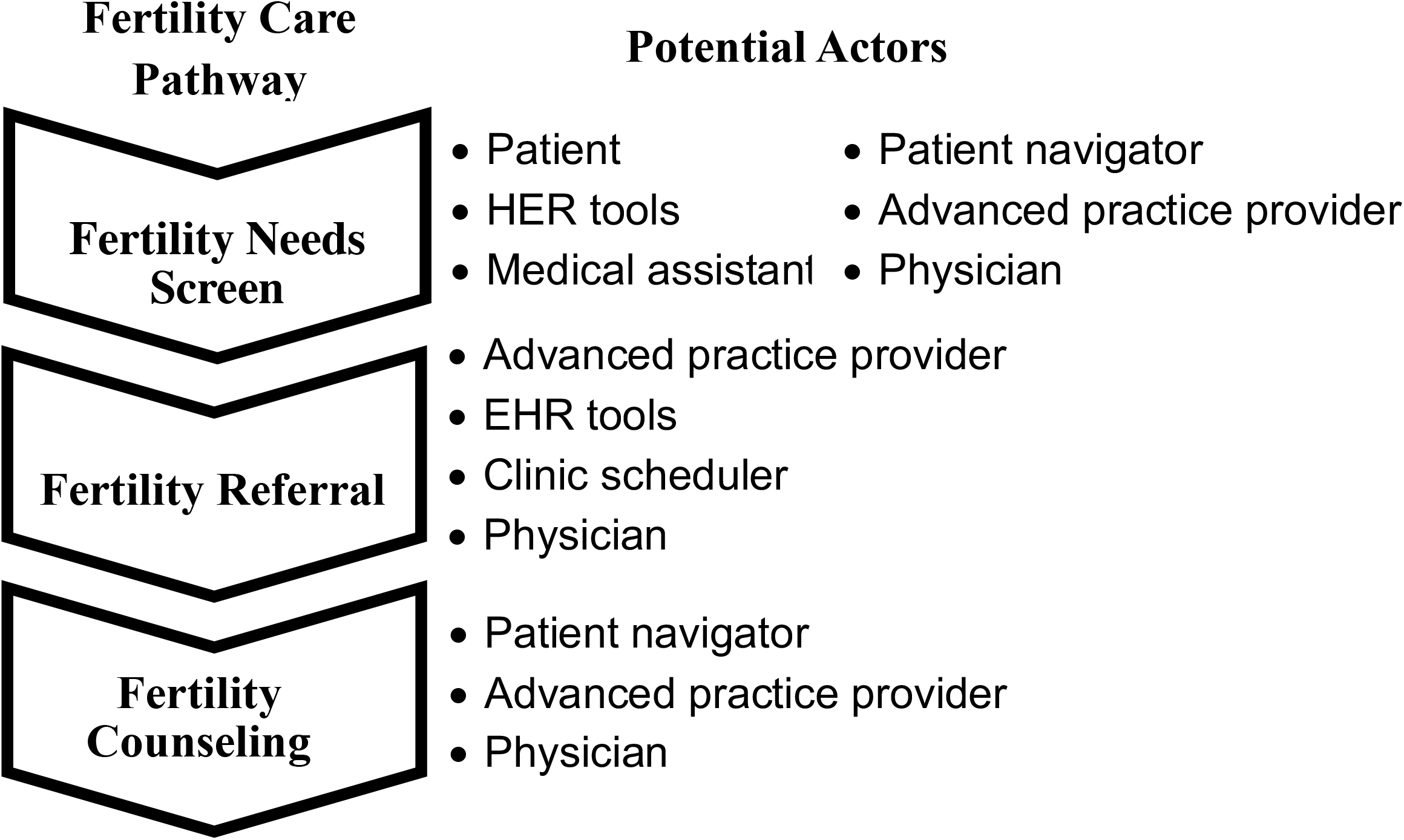
Fertility care pathway involves a fertility care needs screen, a fertility referral, and fertility counseling with multiple potential actors in each step.

### Differences between pediatrics and adult settings

Several themes unique to pediatrics emerged. For adolescents, providers and AYA survivors similarly discussed that the timing of fertility counseling before, concomitant or after consultation with their parents/guardians should be guided by the family. Some providers questioned whether adolescents would be interested in talking about future fertility, believing it was best to talk to the parents only. Pediatric providers also discussed at what age it is appropriate to talk about fertility care: “*Like would the kid want to know about having children at 15 or would you talk just to the parents? I guess you would talk to the parents first and ask do you want to speak about this privately or do you want to speak about it in front of the kid?”* (Advanced practice provider).

These concerns were echoed in the focus groups, as several patients shared that fertility was not on their mind when diagnosed with cancer in adolescence. Additionally, complexity of privacy laws with regard to whether adolescents and/or parents/guardians have access to their patient portal (through which telehealth and screening questionnaires may be delivered) emerged as a barrier to EHR patient portal strategies.

### Mapping implementation strategies to fertility needs screening, referral, and specialist access

Based on barriers and facilitators reported by participants, the investigators designed multiple implementation strategies for the three stages, selected strategies and identified implementors. Figure 3 specifies the five selected strategies for the two oncology programs: 1) automatic fertility needs screen using a best practice advisory (BPA), 2) an opt-out fertility referral pathway through the electronic health record (EHR) system EPIC, 3) adding option to conduct fertility counseling via telehealth, 4) audit and feedback to providers, and 5) conducting educational meetings.

**Figure 3:**
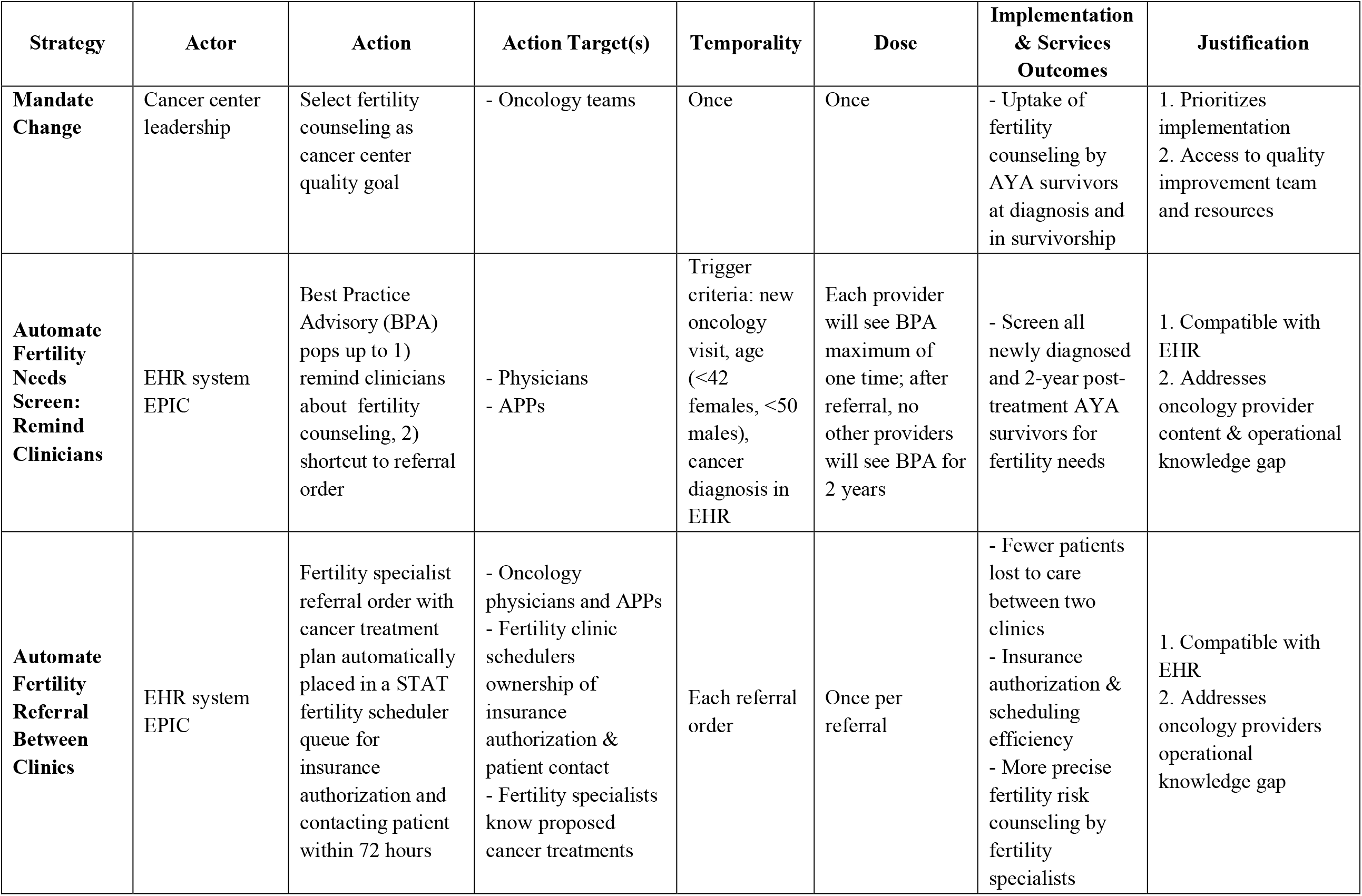
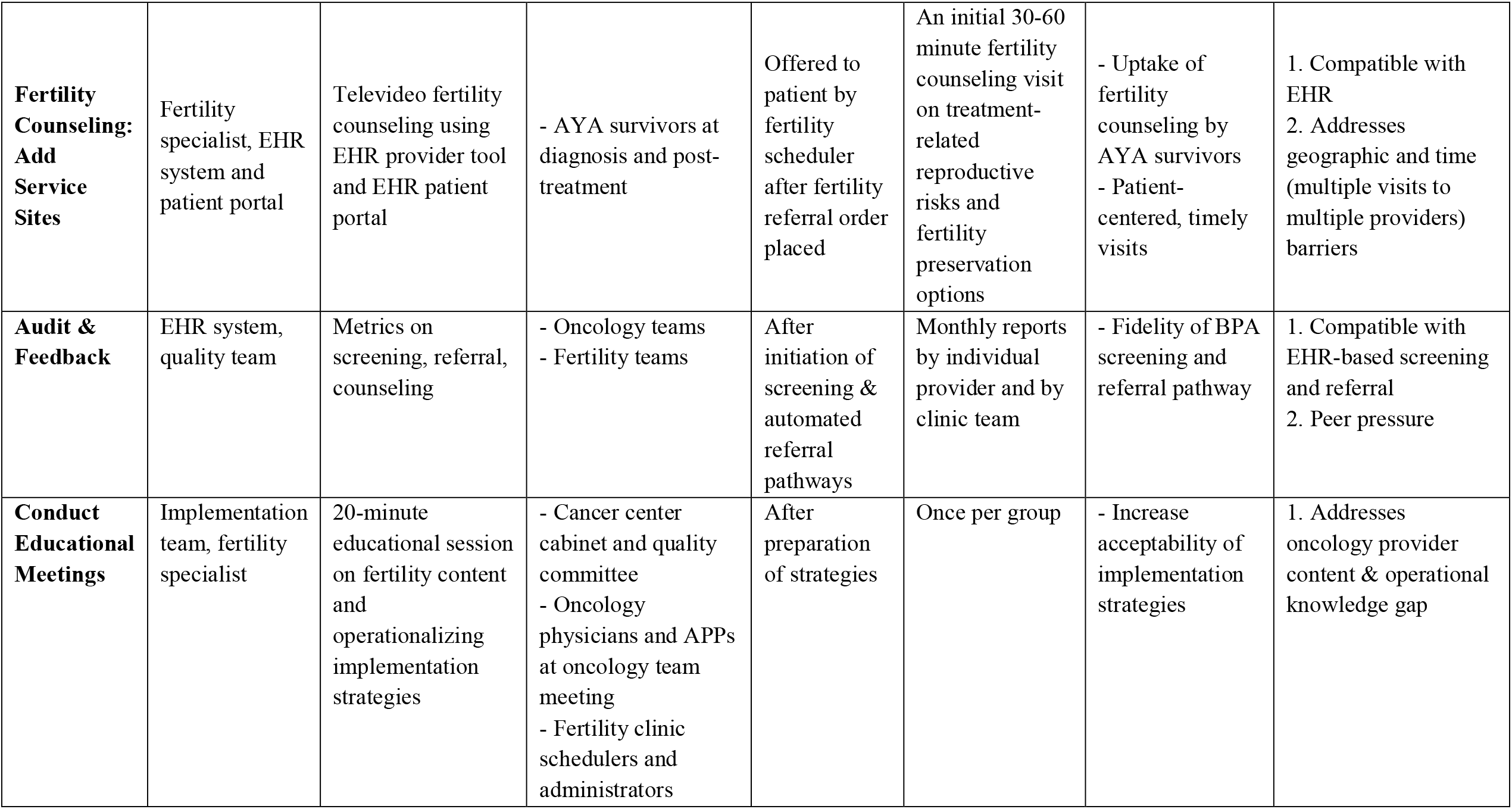
Specifications of selected implementation strategies

The complexity of the BPA, which required significant stakeholder input in planning and EHR programmer expertise in developing, warrants detailing. In the EPIC EHR system, the BPA would pop-up for physicians and advance practice providers at a patient’s first oncology visit based on age (automatic), new visit type (automatic), and that provider’s entering a cancer diagnosis in the encounter. The fertility specialist referral order is defaulted to yes as part of this BPA. Once a provider such as a breast surgeon chooses to refer a patient, no other providers, such as a breast medical oncologist, will see the BPA at any other new-to-clinic visit, to minimize provider fatigue. The next automated BPA appears 2 years later, reasoning that the patient will be post-treatment. If the provider declines to refer, e.g. breast surgeon does not know if the patient has cancer yet, the BPA will pop-up at other providers’ new oncology visits. When providers select not to place the referral, they are required to provide a reason (patient declines to address today, no fertility needs now, poor prognosis, not enough time, already on treatment, other). A priori limitations to the BPA include that uniform criteria for firing will result in deployment at visits when fertility counseling is not appropriate and radiation oncology does not use the same EHR system. Figure 4 specifies other implementation strategies that were considered but not selected.

**Figure 4:**
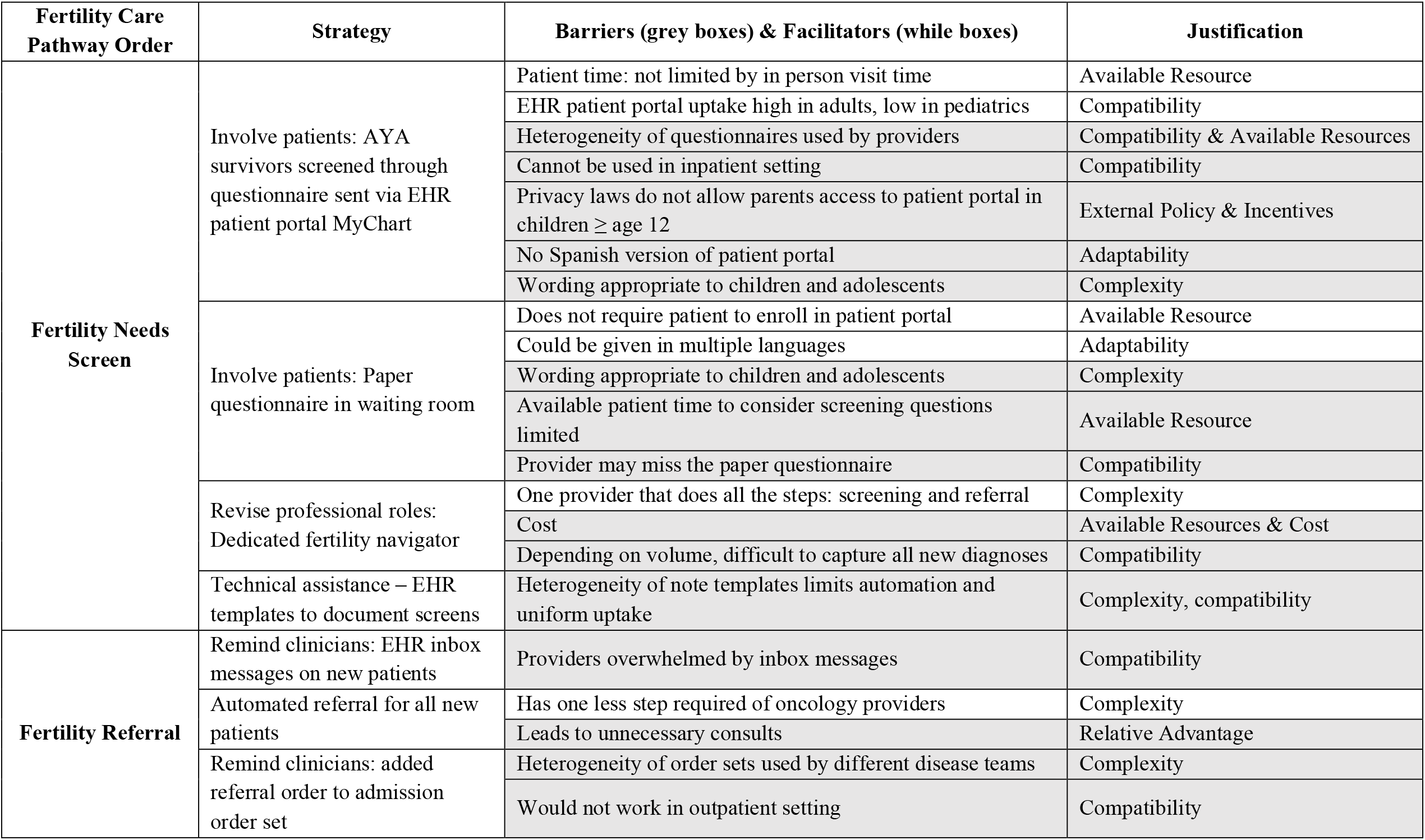
Barriers (grey boxes) and facilitators (white boxes) of non-selected implementation strategies

Key differences between the inpatient and outpatient environment were identified in mapping strategies. EHR tool specifications and pathways require different programming logic between the inpatient and outpatient setting. Inpatient insurance authorizations are not required for consultations, so providers hypothesized that this would improve access to risk counseling by fertility specialists. However, female fertility preservation procedures of oocyte or embryo banking generally occur in outpatient settings, which would limit the access of hospitalized patients.

## Discussion

Guided by an implementation science framework, we conducted a qualitative study with oncology providers and AYA survivors from one adult and one pediatric oncology setting to systematically identify barriers and facilitators to fertility counseling and co-design implementation strategies with stakeholders. We report this systemized approach to inform teams seeking to develop or adapt their implementation of fertility care into routine oncology care.

Using CFIR domains and constructs allowed us to assess systematically the facilitators and barriers that influence fertility counseling implementation and change of processes. CFIR offered a pragmatic structure for our multi-level problem and a large number of domains and constructs that we could query.^23^ While our qualitative guides encompassed questions based on a larger set of constructs, ultimately, the number of key relevant constructs (Figure 1) was smaller and can guide the environmental scan of other clinical settings. Compatibility, feedback, available resources and planning constructs were particularly important in designing specific strategies for the two oncology programs. Recently, we conducted a scoping review of nearly 150 papers on barriers and facilitators to fertility care implementation.^31^ What was striking was the lack of both organized, systematic approaches to identifying factors that influence implementation, as well as how to use these factors to guide selection of implementation strategies, motivating our taking an implementation science approach.

Findings led to conceptualizing three key steps in the implementation of fertility counseling that strategies should target. Then, an array of implementation strategies were considered, taking into account both our findings, compilation of implementation strategies from the Expert Recommendations for Implementing Change project, ^30^ and a priori research on components of fertility care models. ^31^ The goals were to select strategies that fit our context and may be generalizable across adult and pediatric settings; we aimed to minimize the number of strategies to limit complexity and system burden. The process of describing the strategies (naming, defining, operationalizing the actor, action, action targets, temporality, dose, implementation outcomes addressed and theoretical justification) clarified which ones were not feasible (e.g., high quality patient educational materials on fertility risk) or did not fit (e.g., patient portal for screening). ^32^ This reporting may enable scaling fertility counseling implementation efforts.

Across domains, we found variability in beliefs about whose role it was to provide fertility counseling, concordant with prior reports.^14^ For some providers, this concern was due to lack of self-efficacy about addressing fertility, an individual characteristic, while others attributed to inadequate time or incompatibility with a clinical visit, an inner setting characteristic. We also found that many types of providers could perform fertility needs screening, while only physicians and advanced practice providers could place referrals. Hence, specifying the actors of an implementation strategy was necessary.

We sought stakeholder input on EHR tools as implementation strategies. In designing and building EHR-tools, we could modify existing EHR tools such as the reporting tool for audit and feedback and the patient-friendly portal that enables secure video visits. Because of the limited number of EHR software systems, these tools could be widely disseminated and scalable. We also found that EHR tools may not be acceptable due to provider fatigue, feasible due to privacy laws, or appropriate due to low uptake of patient portal apps. Building these tools can be time- and labor-intensive, but a successful build in one clinical program may be disseminated to others.

Limitations require discussion, including that we were limited to two oncology programs. While they were selected to reflect pediatric and adult oncology, their academic and existing fertility clinic referral site limit generalizability. Additionally, our sampling strategy for health care providers and AYA survivors, like other qualitative work, was purposeful and not random. This generates the risk of selection bias as the healthcare providers and AYA survivors who took part in our study may have an interest in fertility care. As we developed implementation strategies, we were limited by available resources. There is a strong need for the creation of educational resources to support fertility risk discussion, where current tools lack specificity and/or are too time intensive. Financial costs are a significant barrier that cannot be overcome with clinic-based implementation strategies alone.

In summary, we describe a systematic approach to design implementation strategies for fertility counseling at an adult and children’s oncology program. We contribute data on the most salient CFIR constructs to assess and specifications on a set of potential strategies that may be adapted and deployed to improve the fertility care of adolescents and young adults with cancer.

## Data Availability

The data that support the findings of this study are available from the corresponding author, [HIS], upon reasonable request.

## Acknowledgments

We wish to thank Kris Brodsho for guidance on EHR capabilities and Nicole Stadnick, PhD for implementation science expertise.

## Notes

**Conflicts of interest:** Dr. Takemoto works for ServiceNow, Inc., which did not sponsor, support, or have oversight of this research.

### Competing Interest Statement

The authors have declared no competing interest.

### Funding Statement

NIH UL1TR001442, NIH TL1TR001443

### Author Declarations

The study was approved by IRBs at University of California, San Diego and Rady Children Hospital

